# Cenobamate as add-on treatment for *SCN8A* developmental and epileptic encephalopathy

**DOI:** 10.1101/2024.10.17.24312949

**Authors:** Cathrine E. Gjerulfsen, Madeleine J. Oudin, Francesca Furia, Sopio Gverdtsiteli, Cecilie Johannessen Landmark, Marina Trivisano, Angel Aledo-Serrano, Ricardo Morcos, Roberto Previtali, Pierangelo Veggiotti, Emilia Ricci, Guido Rubboli, Elena Gardella, Rikke S. Møller

**Affiliations:** Department of Epilepsy Genetics and Personalized Medicine, Danish Epilepsy Center, Dianalund, Denmark; Department of Regional Health Research, Faculty of Health Sciences, University of Southern, Odense, Denmark; Department of Biomedical Engineering, Tufts University, Medford, MA, USA; Department of Pharmacy, Oslo Metropolitan University, Oslo, Norway; The National Center for Epilepsy, Oslo University Hospital, Member of the ERN EpiCare, Oslo, Norway; Department of Pharmacology, Oslo University Hospital, Oslo, Norway; Epilepsy and Movement Disorders Unit, Bambino Gesu Children’s Hospital, IRCCS, Italy; Epilepsy Unit, Vithas Madrid University Hospitals, Madrid, Spain; Universidad Europea de Madrid, Madrid, Spain; Neuroscience Research Center, Department of Biomedical and Clinical Sciences, University of Milan, Milan, Italy; Pediatric Neurology Unit, Buzzi Children’s Hospital, Milan, Italy; Child Neuropsychiatry Unit, Epilepsy Center, San Paolo Hospital, Milan, Italy; Department of Health Sciences, University of Milan, Milan, Italy; Institute of Clinical Medicine, University of Copenhagen, Copenhagen, Denmark; Department of Clinical Neurophysiology, Danish Epilepsy Center, Dianalund, Denmark

**Author notes:** Correspondence to: Professor Rikke S. Møller, M.Sc., PhD, Department of Epilepsy Genetics and Personalized Medicine, Danish Epilepsy Centre, Kolonivej 1, 4293 Dianalund, Denmark.

**Keywords:** Genetic epilepsy, sodium channelopathy, drug-resistant epilepsy, developmental and epileptic encephalopathy

## Abstract

**Objectives:** Developmental and epileptic encephalopathies (DEEs) caused by pathogenic variants in *SCN8A* are associated with difficult-to-treat and early-onset seizures, developmental delay/intellectual disability, impaired quality of life, and increased risk of early mortality. Commonly used antiseizure medications (ASMs) for *SCN8A-*related disorders, caused by gain-of-function variants, are sodium channel blockers. The use of such ASMs is often not enough to gain satisfactory seizure control. In this retrospective study, the effect of cenobamate was assessed in patients with *SCN8A-*DEE.

**Methods:** Across multiple centers and through a collaboration with the patient advocacy organization International *SCN8A* Alliance, patients with *SCN8A*-DEE treated with cenobamate for ≥3 months were identified. Data were obtained once from patients’ caregivers or treating physicians through a RedCap survey. The functional effects of the included patients’ *SCN8A* variants were determined by functional tests published in the literature or functionally classified by prediction tools.

**Results:** Twelve patients (2-25 years, 8 females) with presumed gain-of-function *SCN8A* variants were treated with cenobamate for a mean period of 8.6 months (range 3-27 months). Countable motor seizures were reduced in 12/12 (100%) patients. Seven experienced a seizure reduction above 75% of which two patients achieved seizure freedom. A 25-50% and 50-75% decrease was observed in three and two patients, respectively. An increase in seizure-free days/patient was also reported. Rescue medication was decreased in 83% of patients, and non-seizure-related improvements (increased alertness, better sleep, improved muscle tone) were observed in 58%. Adverse effects were reported by 33%; half resolved spontaneously or by the reduction of concomitant ASMs.

**Significance:** Our data suggest, that cenobamate is a promising and safe treatment for *SCN8A-* DEE, even during early infancy. As a possible precision approach to treatment, cenobamate effectively reduced seizure burden and ameliorated non-seizure-related symptoms. Similar results may be achieved in cohorts of patients with gain-of-function variants encoding other sodium channels.

**Key points:**

- Patients with DEE caused by pathogenic GOF variants in *SCN8A* are commonly treated with sodium channel blockers, often without satisfying seizure control.
- Adjunctive cenobamate improved seizure frequency in 12/12 patients (2-25 years, 8 females) with SCN8A-DEE of which two achieved seizure freedom.
- An increase in seizure-free days/patient, non-seizure-related improvements (e.g. increased alertness), and a decrease in rescue medication was also observed.
- Cenobamate is a promising and safe treatment for *SCN8A-*DEE, even during early infancy.
- Similar results may be achieved in patients with gain-of-function variants encoding other sodium channels.

## Introduction

Pathogenic variants in *SCN8A* encoding the alpha-1 subunit of the Na_V_1.6 sodium channel, have been associated with a range of epilepsies, spanning from self-limiting epilepsy to severe developmental and epileptic encephalopathies (DEEs) (Gardella E 2018, Talwar D 2021, Johannesen KM 2022). A correlation between the functional effects of the variants and the phenotypes has previously been reported, and early-onset DEEs are often caused by gain-of-function (GOF) variants (Johannesen KM 2022). Treatment with sodium channel blockers, especially high doses of phenytoin, carbamazepine, or oxcarbazepine, benefits some affected individuals (Boerma RS 2016, Gardella E 2018, Conecker G 2024, Conecker G 2024). However, seizures in *SCN8A-*related DEEs are often extremely drug-resistant, and only a few patients achieve long periods of seizure freedom (Johannesen KM 2018) (Gardella E 2018).

As one of the newest antiseizure medications (ASMs), cenobamate is developed as a new aryl-carbamate with a dual action mechanism (Löscher W 2021). It is approved as an adjunctive drug for the treatment of focal onset seizures in adults with epilepsy. Cenobamate positively modulates GABA_A_ receptors and blocks persistent sodium currents which increases the inactive state of voltage-gated sodium channels (Löscher W 2021, Roberti R 2021). It has been found effective and safe in randomized, double-blind, placebo-controlled trials (Chung SS 2020, Krauss GL 2020) and long-term Open-Label Extension Studies (Sperling MR 2020, Klein P 2022) in adults, in defiance of drug reaction with eosinophilia and systemic symptoms (DRESS) and low tolerability at high doses may be limitations in its clinical use (Löscher W 2021). In a recent systematic review, cenobamate was evaluated to be the highest-ranked third-generation ASM regarding efficacy (Lattanzi S 2022). Only two studies have reported treating pediatric patients suffering from drug-resistant epilepsy with cenobamate (Makridis KL 2022, Varughese RT 2022), however, the youngest patient in these studies was 10 years old. There are limited reports of treatment with cenobamate in genetic epilepsies, where seizure onset and the development of refractory epilepsy occur in the first few years of life (Makridis KL 2022, Agashe S 2023).

Herein, we present a case series of twelve patients with intractable *SCN8A*-related DEE treated with cenobamate of which one case will be described in detail.

## Methods and patients

We performed a retrospective multicenter study of patients with *SCN8A*-related DEE treated with cenobamate through a collaboration with the patient advocacy organization International *SCN8A* Alliance, and clinicians in Italy and Spain. Clinical information regarding demographics, seizure characteristics and frequencies, treatment details, and outcomes were obtained from patients’ caregivers or treating physicians, who provided anonymized individuals’ data by an online questionnaire (RedCap survey). Data were collected once, and only patients treated for a minimum of three months were included. The functional consequence of the included patients’ *SCN8A* variants was determined by functional tests earlier published in the literature. In case of missing functional results, the variants were functionally classified by prediction tools (SCN viewer, SCION, FunNCion) (Heyne HO 2020, Brunklaus A 2022, Boßelmann CM 2023) in combination with phenotypic features.

In this study, all seizures (focal and generalized) with motor symptoms were grouped as “countable motor seizures”. Non-motor seizures and myoclonic seizures were not included in the analysis due to quantifying difficulties. Seizure types were classified according to the International League Against Epilepsy (Fisher RS 2017). The baseline frequency of countable motor seizures was estimated by the mean value of the countable motor seizures three months before cenobamate initiation. The primary outcome measure was the change in frequency of countable motor seizures, number of seizure-free days, and amount of rescue medication from baseline to data obtained. Additionally, to describe the efficacy of cenobamate on the seizure frequency in a child with *SCN8A* DEE, a case (patient #3) is presented in detail. The secondary endpoints were adjustments of concomitant antiseizure medications (ASMs), non-seizure improvements, adverse effects, and safety.

Caregivers or clinicians provided anonymized individuals’ data via an online questionnaire (Redcap survey). The database was stored at the Danish Epilepsy Centre. Informed consent from the caregivers of the included patients was obtained according to local ethics guidelines. The caregivers of patient #3 consented to a detailed report of the medical history and EEG results in this publication.

## Results

### Cohort demographics

Twelve patients with *de novo SCN8A* missense variants (aged 2-25 years, 8 females) were included in the analysis. One individual (patient #3, supplementary tale 1) harbored two missense variants in cis p.(Thr144Ser) and p.(Ser217Pro). Both variants have been functionally characterized and p.(Ser217Pro) is causing a clear gain-of-function (GOF) whereas p.(Thr144Ser) did not show any functional effect (Vanoye CG 2024). Two additional variants have previously been functionally characterized and shown to cause a GOF (patients #1 and #12, supplementary table 2) (Wagnon JL 2015, Liu Y 2019). The remaining eight *SCN8A* variants were predicted to cause GOF based on the functional characterization of the paralogue variants in other sodium channel genes and prediction models combined with phenotypic features (supplementary table 2). The majority of patients presented with global developmental delay (11/12, 92%) and intellectual disability (9/12, 75%). Three patients were too young to determine the grade of intellectual abilities. The mean onset of seizures was 3.6 months (range 0-12 months) (supplementary table 1). At baseline, 9/12 (75%) patients had a frequency of countable motor seizures (tonic, generalized tonic-clonic, focal to bilateral tonic-clonic, and focal motor seizures) higher than 20 per month, and 11/12 (92%) were treated with at least two concomitant ASMs (table 1). One patient did not receive treatment with concomitant ASMs at baseline (patient #7, supplementary table 1). The most commonly concomitant ASMs were lacosamide, clobazam, clonazepam, oxcarbazepine, and phenytoin. At baseline, 3/12 (25%) were treated with ketogenic diet, and 2/12 (17%) had a Vagus Nerve Stimulator (VNS). Comorbidities were reported in 12/12 (100%) of the patients consisting of severe hypotonia, cortical visual impairments, feeding difficulties, incontinence, movement disorders (chorea, dyskinesia), neuropsychiatric features (ADHD, aggressiveness), hearing loss, and sleep disorders (sleep apnea, insomnia).

**Table 1:**
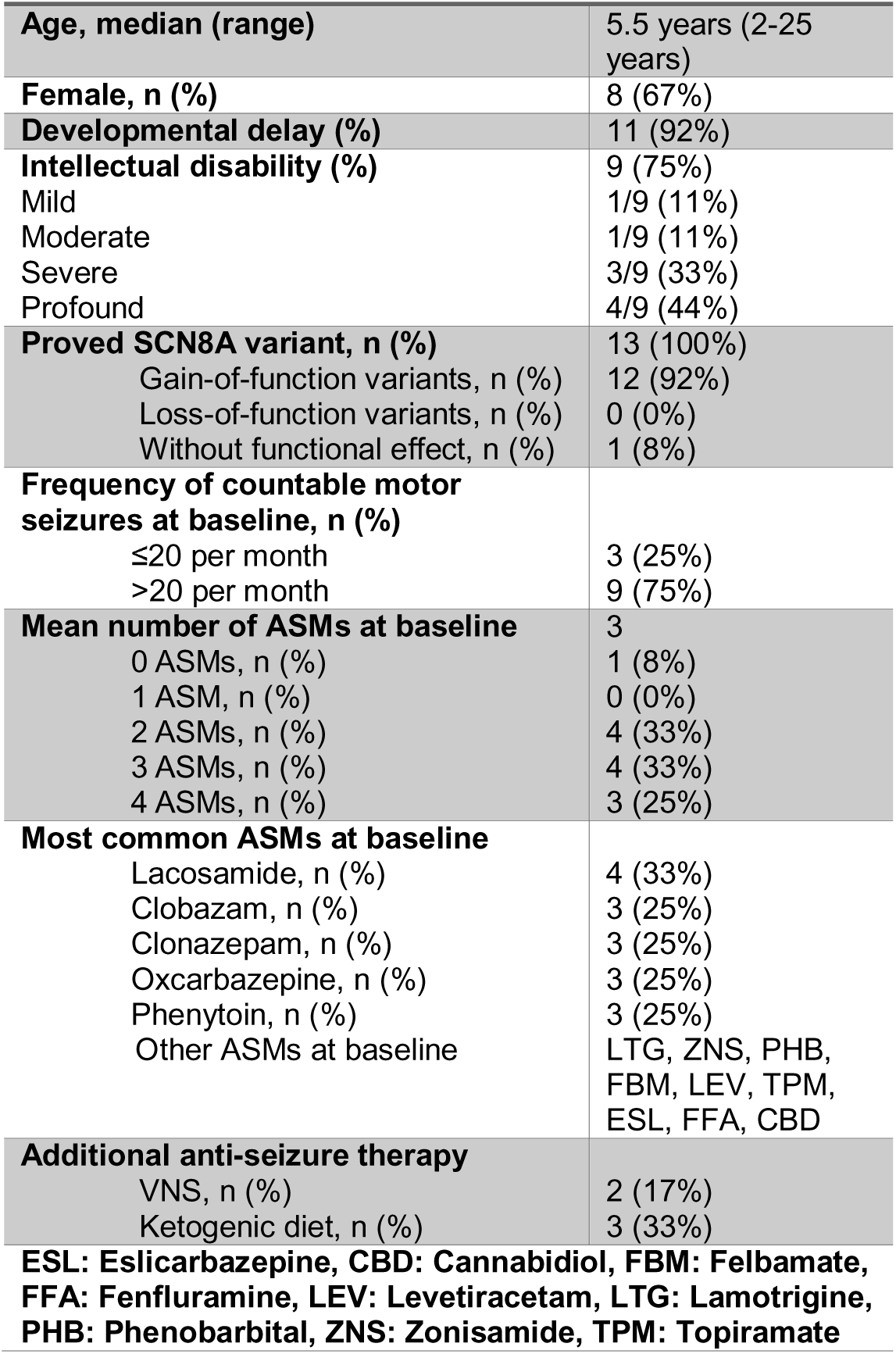
Baseline characteristics (n = 12) Baseline characteristics of the12 included patients with *SCN8A-*DEE treated with cenobamate.

### Cenobamate dose, seizure reduction, and concomitant treatment

All patients were treated with cenobamate for at least three months including titration, and the mean length of treatment of the whole cohort was 8.6 months. The titration scheme applied for the cohort varied according to the differences in patients’ weight (table 2).

**Table 2:** Cenobamate treatment and seizure outcomes at data collection. The distribution of seizure outcomes of the 12 treated patients in relation to baseline seizure frequency, maintenance dose of cenobamate, and length of treatment period.

| **Table 2: Cenobamate treatment and seizure outcomes at data collection** | **100% reduction** | **99-75% reduction** | **74-50% reduction** | **49-20% reduction** |
| --- | --- | --- | --- | --- |
| **Total cohort n, (%)** | 2 (17%) | 5 (42%) | 2 (17%) | 3 (25%) |
| **Freq. of seizures at baseline, n (%)** |  |  |  |  |
| ≤20 per month | 0 | 5 (100%) | 1 (50%) | 2 (66%) |
| >20 per month | 2 (100%) | 0 | 1 (50%) | 1 (33%) |
| **Cenobamate maintenance dose, mean (range)** | 50mg (50mg) | 135mg (62.5-250mg) | 100mg (100mg) | 112.5mg (50-200mg) |
| **Length of treatment period** |  |  |  |  |
| <6 months (n=3) | 0 | 1 (20%) | 0 | 2 (67%) |
| ≥6months and <1 year (n=6) | 2 (100%) | 1 (20%) | 2 (100%) | 1 (33%) |
| ≥1 year (n=3) | 0 | 3 (60%) | 0 | 0 |

The majority of the patients were below the age of 18 years (10/12, 83%). In the pediatric patients, the mean initial dose of cenobamate was 0.46 mg/kg/day (0.25-0.74mg/kg/day) with a mean body weight of 18.8 kg (13-29.5 kg), and the maximum dose given was 250mg. The two adult patients (mean age 22 years and mean body weight 45.4 kg) followed the titration scheme recommended by the official guidelines (SK Life Science 2021), and the maximum dose of cenobamate given was 200mg.

A reduction in the frequency of countable motor seizures was observed in all the patients (12/12, 100%) after the introduction of cenobamate (supplementary table 1, Figure 1). A seizure reduction of 25-50% and 50-75% were observed in three and two patients respectively. Additionally, seven individuals experienced a seizure reduction above 75% of which two patients, who also share the same variant of *SCN8A,* reached a reduction of countable motor seizures at 100% (patients #8 and#11).

**Figure 1:**
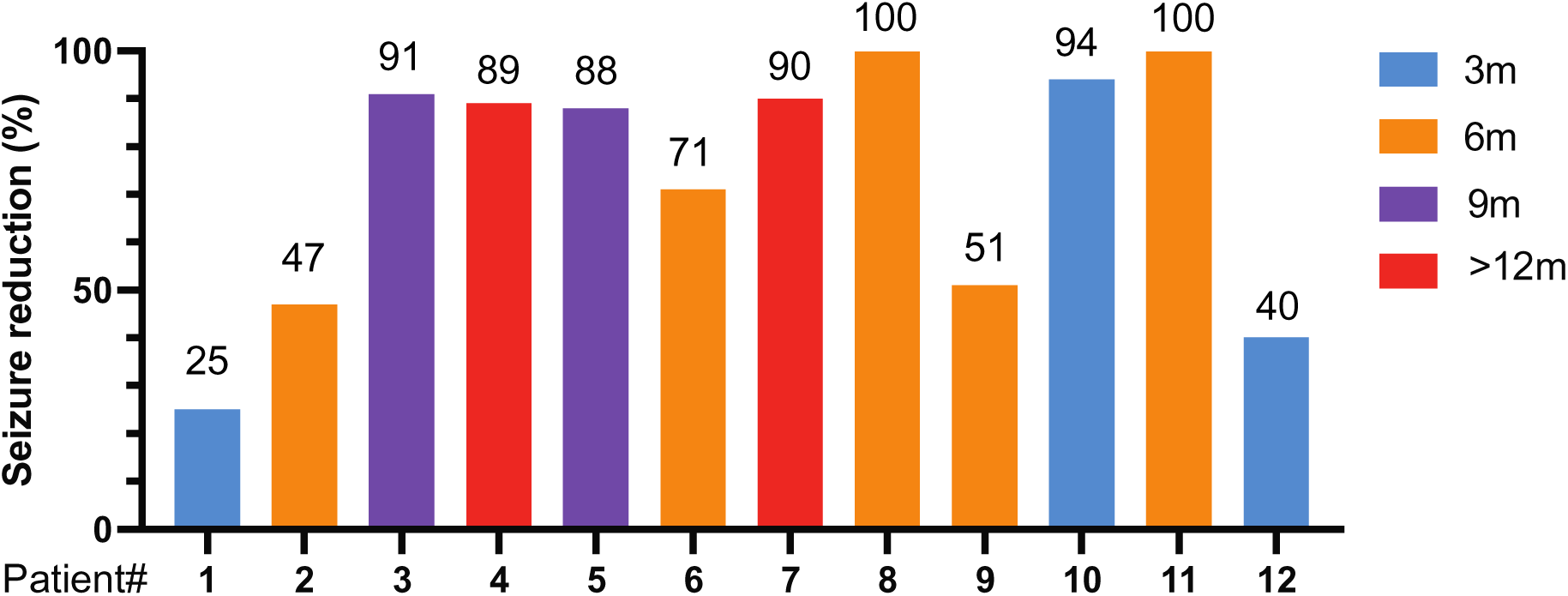
The percentage of seizure reduction for each patient with follow-up at 3 months, 6 months, 9 months, or >12 months.

As visualized in table 2, the seizure frequency decreased with the length of treatment. However, the cenobamate maintenance dose and frequency of seizures at baseline did not seem to influence the outcome.

At baseline, 7/12 (58%) patients suffered from daily seizures. After adjunctive treatment with cenobamate, the number of patients having monthly seizure-free days increased from 5/12 (42%) to 9/12 (75%). Of the 42% of the patients having seizure-free days at baseline, 4/5 (80%) experienced an increase in the number of seizure-free days after the initiation of cenobamate.

Information on the use of rescue medication was available in 9/12 patients, of which 5/9 (56%) commonly used rescue medication at baseline. A reduction in the administration of rescue medication after the initiation of cenobamate was observed in 5/6 (83%) individuals (supplementary table 1). Additionally, non-seizure improvements consisting of increased alertness, better sleep, more wakeful hours of the day, increased vocabulary, and improved muscle tone were reported by parents and clinicians in 7/12 (58%) patients.

In 9/12 patients, concomitant ASMs were adjusted after the initiation of cenobamate of which the most often observed changes were reductions of clobazam and sodium channel blockers (oxcarbazepine, lacosamide, phenytoin) either proactively, as has been recommended (Smith MC 2022), or due to adverse effects (somnolence, increased hypotonia, or reduced alertness) caused by drug-drug interactions.

### Safety and tolerability

In the present cohort, 4/12 (33%) patients reported adverse effects (e.g. somnolence, constipation, worsening of hypotonia, reduced alertness) related to cenobamate treatment of which two resolved (patients #3 and #10, supplementary table 1) either spontaneously or by the reduction of concomitant ASMs (clobazam and phenytoin). The adverse effects reported in patients #2 and #4 (somnolence) were due to drug-drug interactions between clobazam and cenobamate; no further data after this adjustment is available. No patients discontinued cenobamate and no serious adverse events (eg. DRESS) were observed.

### Case presentation

As a drug approved for the treatment of focal onset seizures in adults with epilepsy, cenobamate is not the first ASM selected when treating children with refractory epilepsy and several comorbidities. In the literature, the youngest patient treated with cenobamate is 10 years of age. In this study, we report the use of cenobamate in a cohort of patients of which 8/12 patients were between the ages of 2 and 10 years old. As the reported results visualize, children with DEE may benefit from adjunctive treatment with cenobamate. The following case describes the use of cenobamate in suffering from *SCN8A*-DEE.

Patient #3 (supplementary table 1) is the first child, born by non-consanguineous parents without a family history of epilepsy. The pregnancy was uncomplicated, and the development was normal until seizure onset, which was followed by severe developmental delay. Electroencephalography (EEG) showed multifocal discharges, and MRI was normal. Exome sequencing revealed two *de novo* missense variants (c.649T>C, p.(Ser217Pro), and c.431C>G, p.(Thr144Ser)) in *SCN8A*. The patient was then diagnosed with infantile epileptic spasm syndrome, which recurred in spite of prednisolone, vigabatrin and ACTH treatment, and were eventually controlled with the ketogenic diet. After, she has continued to suffer from clonic and tonic seizures with an average frequency of 33 seizures/day in total, in spite of treatment with levetiracetam, clobazam, oxcarbazepine, lacosamide, phenytoin, cannabidiol, and valproic acid. In addition to refractory epilepsy, the patient is diagnosed with cortical visual impairment, hypotonia, congenital laryngomalacia, and bilateral intermittent exotropia.

The patient started treatment with cenobamate 6.25mg every other day in combination with clobazam, levetiracetam, and phenytoin (figure 2, B). Cenobamate was slowly titrated to 75mg once daily over six months, and the frequency of tonic seizures was reduced to 3 seizures per day. Additionally, clonic seizures almost completely disappeared. The interictal EEG showed very frequent multifocal epileptiform abnormalities at baseline, that were significantly reduced after cenobamate treatment (Figure 2 C-D). Clobazam was decreased from 15mg/day to 5 mg/day and phenytoin from 150 mg/day to 87.5mg/day, but the dose of levetiracetam was unchanged.

**Figure 2:**
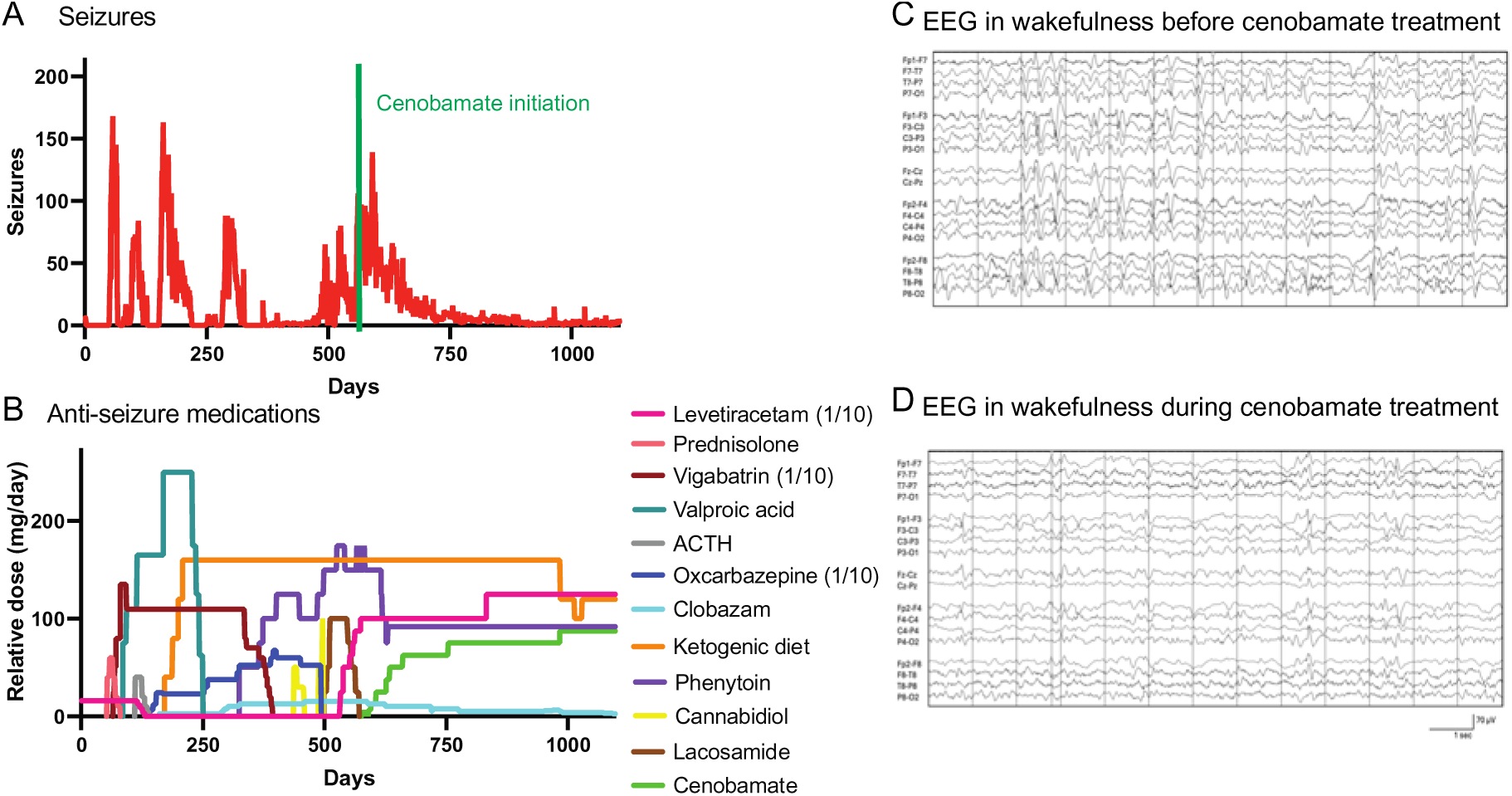
Seizure frequency (A) and antiseizure medication (B) from of patient #3, described in detail. EEG results from before and after cenobamate treatment (C and D).

## Discussion

To our knowledge, this is the first report of patients with *SCN8A*-DEE treated with cenobamate, with a median patient age of only 5.5 years old. We found that cenobamate was safe and effective in all patients despite severe refractory epilepsy and severe health conditions complicated by several comorbidities.

### Efficacy and tolerability

By adjunctive treatment with cenobamate, all individuals in our study achieved clinically meaningful improvement in the frequency of countable motor seizures with a median seizure reduction of 88.5% overall and a retention rate at the last follow-up of 100%. Three patients had only been treated for three months each and reached a median seizure reduction of 40%. It is possible, that the median seizure reduction of the total cohort will increase with the treatment period, as the first three months of treatment also include titration. A reduction of rescue medication in 5/6 (83%) patients was also reported as well as non-seizure-related improvements such as increased alertness, better sleep, increased vocabulary, and improved muscle tone.

A case presentation was included to better describe the *SCN8A*-DEE phenotype and the benefits of treatment with cenobamate. After treatment with 11 different ASMs and a persistent high seizure burden, adjunctive therapy with cenobamate decreased the frequency of countable motor seizures by 91%. Like most of the included patients in our cohort, the patient did not experience permanent adverse effects. However, a discussed limitation in its use may be the risk of DRESS, short QT-syndrome, and low tolerability at high doses, and therefore studies evaluating clinical experience are of great importance (Löscher W 2021, Steinhoff BJ 2024). In our cohort, the patients had limited language or were non-verbal, which may have impeded the report of adverse effects. Nevertheless, most patients were cared for by parents or established caregivers, who generally can observe even small changes in the patients’ clinical condition.

There are pharmacological challenges with cenobamate, as it has considerable variability in dose and exposures within and between patients, which makes individual dosing schemes unpredictable (Steinhoff BJ 2024). Also, cenobamate acts as a combined enzyme inducer and inhibitor, which may alter the serum concentrations considerably and unpredictably (Johannessen Landmark C 2023). An example is variable and up to several-fold increase in the active metabolite of clobazam, desmethyl-clobazam, which may cause excessive adverse effects (Elakkary S 2023). This elucidates the need for therapeutic drug monitoring when risk of pharmacokinetic interactions in complex therapeutic regimes.

### Quality of life evaluation

As this study was performed retrospectively, it was not possible to evaluate if treatment with cenobamate consistently increased the quality of life (QoL) for patients or caregivers. Patients with *SCN8A-*DEE are burdened by high seizure frequencies and multiple comorbidities, which are known to decrease patients’ and caregivers’ QoL in general (Auvin S 2021). As observed in patients with other DEEs, seizure-free days and reduced seizure frequency increase patients’ and caregivers’ QoL overall. Additionally, it has been found that seizure-free days have a greater impact on QoL than seizure frequency in total (Auvin S 2021). In our cohort, the number of patients experiencing monthly seizure-free days after the initiation of cenobamate increased to 75%. Additionally, 80% of the patients having sporadic seizure-free days at baseline, experienced an increase in the number of seizure-free days after the initiation of cenobamate. Thus, we may speculate, that cenobamate most likely increased the QoL in our cohort’s patients and caregivers.

### Relation between mechanism of action and genetic variants in SCN8A

With its dual action mechanism, cenobamate modulates GABA_A_ receptors and blocks persistent sodium currents, which distinguishes it from other sodium channel blockers like carbamazepine and lamotrigine, that target the transient sodium channel current (Roberti R 2021). Earlier, persistent sodium current has been suggested to be important in normal neuronal excitability and a target for effective seizure control (George 2004, Anderson LL 2014). The included patients all harbored presumed GOF variants and were mainly treated with sodium channel blockers in combination with other ASMs. The improvement of seizure control after the introduction of cenobamate supports a beneficial effect of the blockage of persistent sodium current. As cenobamate also binds at GABA_A_ -receptors at a non-benzodiazepine binding site, which causes hyperpolarization and thus increases the excitation threshold. It is unclear whether the combination of the two distinguished mechanisms of action or a yet undescribed mechanism of cenobamate is the reason for the efficacy (Löscher W 2021) specifically in patients with *SCN8A*-DEE. Thus, this may elucidate a precision approach to the treatment of this severe DEE. In addition, results similar to those observed in our cohort may also be achieved in cohorts of patients with *SCN1A* and *SCN2A* GOF variants and not exclusive for patients with *SCN8A* variants. Adjunctive treatment with cenobamate has also been tried in adult patients with Dravet syndrome caused by *SCN1A* loss-of-function variants (LOF) of which four out of four treated patients achieved a seizure reduction at 80-90% (Makridis KL 2022). Treatment with sodium channel blockers in patients with *SCN1A* LOF variants is normally associated with seizure aggregation and a decrease in cognitive abilities (de Lange IM 2018). The favorable response in the reported patients with Dravet syndrome treated with cenobamate may be due to shared LOF and GOF properties of the variants, or abnormal neuronal networks caused by prolonged disease with high seizure burden. However, this and our report indicate that the potential of cenobamate may be broader than limited to adult patients with drug-resistant focal epilepsy. Adjunctive treatment with cenobamate should therefore be considered when treating refractory genetic epilepsy syndromes also below the age of 10 years of age.

### Methodological considerations

Some limitations of this study must be considered. First, as a retrospective and non-interventional study, the treatment with cenobamate was neither randomized nor blinded to patients’ caregivers or physicians. Secondly, only motor seizures were counted due to difficulties in quantifying other seizure types. Therefore, actual seizure frequencies may have been underestimated. Third, serum concentration measurements were not yet available for cenobamate; thus, pharmacokinetic variability was not investigated. The small sample size is also a limitation of the study. However, *SCN8A-*DEE is a rare disease, and cenobamate is only registered for treating focal epilepsy in adult patients, which limits the number of patients in treatment. The inclusion of patients with other variants of genes encoding sodium channels may further increase our knowledge about the treatment with cenobamate.

In conclusion, our results suggests that adjunctive treatment with cenobamate is an effective and safe treatment option for patients with *SCN8A-*DEE and potentially other DEEs of genetic etiology including patients with sodium channelopathies as a precision approach.

## Acknowledgments

The authors thank the patients and families who participated in this study. The case series was implemented in collaboration with the *SCN8A* Alliance in the USA, which contributed greatly to the collection of data.

## Data availability statement

The data that support the findings of this study are available on request from the corresponding author. The data are not publicly available due to privacy or ethical restrictions. Most of the data that supports the findings of this study are available in the supplementary material of this article

## Ethical publication statement

We confirm that we have read the Journal’s position on issues involved in ethical publication and affirm that this report is consistent with those guidelines.

## Supplementary data

**Supplementary Table 1.** A summary of patients’ important clinical characteristics, cenobamate treatment, and outcome.

| # | SCN8A Variant | Gender / Age (y) | Sz onset | DD / ID status (grade) | Treatment period | CNB dose initiated | CNB maintenance dose | Concomitant ASMs | Freq. of countable motor sz at baseline | Reduction in freq. of countable motor sz | Change in amount of rescue medication | Adverse effects |
| --- | --- | --- | --- | --- | --- | --- | --- | --- | --- | --- | --- | --- |
| 1 | c.5615G>T, p.(Arg1872Leu) | F / 21-25y | 0-3m | Yes / Yes (profound) | 3m | 12.5mg (0.35mg/kg/day) | 200mg (5.6mg/kg/day) | CLB, OXC, PHB, FBM, (VNS) | 89 sz/m | 25% | Baseline: 41 doses<br>3m after: 33 doses | None |
| 2 | c.2620G>A, p.(Ala874Thr) | F / 16-20y | 7-12m | Yes / Yes (severe) | 6m | 12.5mg (0.23mg/kg/day) | 87.5mg (1.6mg/kg/day) | CLB, ZNS, FFA | 17 sz/m | 47% | None | Somnolence (in comb. with CLB) |
| 3 | c.649T>C, p.(Ser217Pro) and c.431C>G, p.(Thr144Ser) | F / 0-5y | 0-3m | Yes / NA | 16m | 6.25mg every 2<sup>nd</sup> day (0.5mg/kg every 2<sup>nd</sup> day) | 75mg (6.25mg/kg/day) | CLB, LEV, PHE, (KetoDiet) | 990 sz/m | 91% | Baseline: 8 doses<br>9m after: 2 doses | Constipation (resolved) |
| 4 | c.718 A>C, p.(Ile240Leu) | F / 11-15y | 0-3m | Yes / Yes (profound) | 2y 3m | 12.5mg (0.4mg/kg/day) | 212.5mg (8.7mg/kg/day) | LCS, DZP, (KetoDiet) | 36 sz /m | 89% | None | Hypotonia, somnolence (in comb. with CLB) |
| 5 | c.2603T>C, p.(Ile868Thr) | F / 6-10y | 0-3m | Yes / Yes (profound) | 9m | 6.25mg (0.28mg/kg/day) | 250mg (11.4mg/kg/day) | CBZ, OXC, (VNS) | 300 sz/m | 88% | NA | None |
| 6 | c.802A>C, p.(Ile268Leu) | F / 11-15y | 4-6m | Yes / Yes (mild) | 6m | 6.25mg (0.22mg/kg/day) | 100mg (3.5mg/kg/day) | ESL, LMT | 7 sz/m | 71% | Baseline: 3 doses<br>5m after: 1 dose | None |
| 7 | c.1790G>A, p.(Arg597His) | M / 0-5y | 4-6m | Yes / Yes (severe) | 13m | 12.5mg (0.65mg/kg/day) | 75mg (3.9 mg/kg/day) | - | 600 sz/m | 90% | None | None |
| 8 | c.727G>T, | M / 0-5y | 4-6m | Yes / NA | 6m | 6.25mg | 50mg (3.2 | CLB, LCS, | 300 sz/m | 100% | None | None |
|  | p.(Ala243Ser) |  |  |  |  | (0.4mg/kg/day) | mg/kg/day | TPM |  |  |  |  |
| 9 | c.2620G>A,<br>p.(Ala874Thr) | F / 0-5y | 4-6m | Yes / Yes<br>(severe) | 6m | 12.5mg<br>(0.7mg/kg/day) | 100mg (5.6<br>mg/kg/day) | CBD CNZ,<br>LCS, FFA | 43 sz/m | 51% | Baseline: 4 doses<br>4m after: NA | None |
| 10 | c.4410A>C,<br>p.(Gln1470His<br>) | F / 6-10y | 0-3m | Yes / Yes<br>(moderate<br>) | 3m | 12.5mg<br>(0.74mg/kg/day) | 62.5mg<br>(0.27mg/kg/<br>døgn) | CBZ, CNZ,<br>PHT | 32 sz/m | 94% | Baseline: >12<br>doses<br>3m after: 3 doses | Reduced<br>alertness<br>(resolved) |
| 11 | c.727G>T,<br>p.(Ala243Ser) | M / 0-5y | 0-3m | No / NA | 6m | 6.25mg<br>(0.45mg/kg/day) | 50mg (NA) | CNZ, LCS,<br>PHE, TPM | 30 sz/m | 100% | NA | None |
| 12 | c.5614C>T;<br>p.Arg1872Trp | M / 0-5y | 4-6m | Yes / Yes<br>(profound<br>) | 3m | 6.25mg<br>(0.48mg/kg/day) | 50mg<br>(3,8mg/kg/d<br>ay) | OXC, LMT,<br>(KetoDiet) | 5 sz/m | 40% | Baseline: 2 doses<br>3m after: 2 doses | None |

CLB: Clobazam, CBD: Cannabidiol, CBZ: Carbamazepine, CNZ: Clonazepam, DZP: Diazepam, ESL: Eslicarbazepine, FBM: Felbamate, FFA: Fenfluramine, KetoDiet: Ketogenic diet, LEV: Levetiracetam, LCS: Lacosamide, LMT: Lamotrigine, OXC: Oxcarbazepine, PHE, Phenytoin, TPM: Topiramate, VNS: Vagus nerve stimulator
DD: Developmental delay, Freq.: Frequency, ID: Intellectual disability, M: Month, Sz: Seizure, Y: year

**Supplementary table 2.** The functional effect of the SCN8A variants determined by functional test earlier published in the literature and estimated by prediction tools.

| No . | SCN8A Variant | Functional study result (ref) | SCN-viewer (ref) | funNCion prediction | SCION prediction |
| --- | --- | --- | --- | --- | --- |
| 1 | p.(Arg1872Leu) | GOF (Wagon, 2015) | GOF | GOF 83% | GOF 100% |
| 2 | p.(Ala874Thr) |  | Unknown | GOF 69% | GOF 70% |
| 3 | p.(Ser217Pro) | GOF (Vanoye, 2024) | GOF in SCN9A paralogue with same AA change (Estacion, 2010) | Unreliable prediction (GOF 60%) | GOF 71% |
|  | p.(Thr144Ser) | No effect (Vanoye, 2024) | Unknown | Unreliable prediction (LOF 75%) | GOF 62% |
| 4 | p.(Ile240Leu) |  | GOF in SCN9A paralogue with different AA change (Ahn, 2010) | GOF 88% | GOF 83% |
| 5 | p.(Ile868Thr) |  | GOF in SCN3A and SCN9A paralogues with same AA change (Zaman, 2018; Zaman, 2020, Cummins, 2004; Namer, 2015; Theile, 2011) | Unreliable prediction (GOF 85%) | GOF 80% |
| 6 | p.(Ile268Leu) |  | Unknown | GOF 89% | GOF 78% |
| 7 | p.(Arg597His) |  | Unknown | Unreliable prediction (LOF 59%) | GOF 67% |
| 8 | p.(Ala243Ser) |  | Unknown | GOF 59% | GOF 56% |
| 9 | p.(Ala874Thr) |  | Unknown | GOF 69% | GOF 70% |
| 10 | p.(Gln1470His) |  | GOF in SCN1A paralogue with same AA change (Barbieri, 2019; Moreau, 2013) | GOF 80% | GOF 79% |
| **11** | p.(Ala243Ser) |  | Unknown | GOF 59% | GOF 56% |
| **12** | p.(Arg1872Trp<br>) | GOF (Liu, 2019) | GOF | GOF 76% | GOF<br>100% |

